# A Rapid, Portable, and Electricity-Free Sample Extraction Method for Enhanced Molecular Diagnostics in Resource-Limited Settings

**DOI:** 10.1101/2023.11.06.23298007

**Authors:** Ivana Pennisi, Mathew L. Cavuto, Luca Miglietta, Kenny Malpartida-Cardenans, Oliver W Stringer, Katerina-Theresa Mantikas, Ruth Reid, Rebecca Frise, Nicolas Moser, Paul Randell, Frances Davies, Frances Bolt, Wendy Barclay, Alison Holmes, Pantelis Georgiou, Jesus Rodriguez-Manzano

**Author notes:** **Corresponding Author Jesus Rodriguez-Manzano** − Department of Infectious Disease, Faculty of Medicine, Imperial College London, UK. **Email:**. These authors contributed equally to the work (Ivana Pennisi, Mathew L. Cavuto).

## Abstract

The COVID-19 pandemic has highlighted the need for rapid and reliable diagnostics that are accessible in resource-limited settings. To address this pressing issue, we have developed a rapid, portable, and electricity-free method for extracting nucleic acids from respiratory swabs (i.e.nasal, nasopharyngeal and buccal swabs),successfully demonstrating its effectiveness for the detection of SARS-CoV-2 in residual clinical specimens. Unlike traditional approaches, our solution eliminates the need for micropipettes or electrical equipment, making it user-friendly and requiring little to no training. Our method builds upon the principles of magnetic bead extraction and revolves around a low-cost plastic magnetic lid, called SmartLid, in combination with a simple disposable kit containing all required reagents conveniently pre-aliquoted. Here, we clinically validated the SmartLid sample preparation method in comparison to the gold standard QIAamp Viral RNA Mini Kit from QIAGEN, using 406 clinical isolates, including 161 SARS-CoV-2 positives, using the SARS-CoV-2 RT-qPCR assays developed by the US Centers for Disease Control and Prevention (CDC). The SmartLid method showed an overall sensitivity of 95.03% (95% CI: 90.44%–97.83%) and a specificity of 99.59% (95% CI: 97.76%–99.99%), with a positive agreement of 97.79% (95% CI: 95.84%–98.98%) when compared to QIAGEN’s column-based extraction method. There are clear benefits to using the SmartLid sample preparation kit: it enables swift extraction of viral nucleic acids, taking less than 5 minutes, without sacrificing significant accuracy when compared against more expensive and time-consuming alternatives currently available on the market. Moreover, its simplicity makes it particularly well-suited for the point-of-care where rapid results and portability are crucial. By providing an efficient and accessible means of nucleic acid extraction, our approach aims to introduce a step-change in diagnostic capabilities for resource-limited settings.

## INTRODUCTION

Seasonal respiratory infections pose a significant public health problem due to their high rate of transmission and wide variety of symptoms^1^. As it is often challenging to identify the precise virus causing a respiratory infection based solely on symptoms, timely and accurate diagnostic testing is critical to successfully containing and treating infection outbreaks^2^. If viral outbreaks are allowed to reach pandemic-level prevalence, as witnessed with the COVID-19 pandemic, economies and communities can be severely impacted and face long-lasting damage^3,4,5^.

Current diagnostic methodologies typically rely on laboratory approaches that identify pathogens or monitor antibody levels in bodily fluids^6^. The most used techniques involve the identification of nucleic acids or proteins. While protein-based immunoassays, such as lateral flow devices, are useful tools for estimating overall population infection rates, they often lack enough sensitivity to reliably identify patients with low viral load, particularly during early and asymptomatic stages of infection^7,8^.Furthermore, such tests are prone to cross-reacting non-specifically with antibodies of other related viruses, limiting their ability to distinguish true positives from false positives^9^. Thus, because of their superior sensitivity and specificity, molecular nucleic acid-based detection methods, such as real-time PCR, have become the gold standard for viral infection diagnosis^10,11,12^.

However, such molecular detection methods typically entail complicated laboratory techniques, require specialized equipment, and rely on extensive technical knowledge and training of operators. Additionally, necessary preanalytical components (e.g., sample collection, transportation, preparation, etc.),contribute to a slow turnaround time; further delaying findings and related diagnostic, as well as therapeuticdecisions^13^. Of these preanalytical components, sample preparation and extraction in particular contribute to a significant portion of the sample-to-result time. This is because the sensitivity of PCR, and amplification chemistries in general, are highly dependent on the purity and concentration of the extracted nucleic acid sample, leading often to lengthy and complex sample preparation protocols^14–16^.

Due to these requirements and complexities, access to molecular methodologies can be restricted, especially in resource limited settings where less accurate immunoassays are often employed^17^. Therefore, there is a clear and pressing need to develop alternative and more accessible diagnostic technologies^18,19^.Since the advent of the 2020 pandemic, multiple laboratories have developed novel alternative approaches, including rapid and simple sample preparation platforms that are less reliant on strained supply chains. However, these improved approaches still require significant use of bench top electrical equipment, trained technical staff, and a controlled laboratory environment^20, 21^.

To overcome these limitations, our group has developed a novel purification method and kit, called SmartLid that includes everything required for a rapid and high-quality nucleic acid extraction in a simple power-free kit. SmartLid is designed to work with a variety of readily available consumable tubes, making it adaptable to any setting and environment.

The kit utilizes a custom magnetic lid and magnetic nanoparticles (or beads) to transfer targeted nucleic acids through three simple sample processing steps, making it a rapid and cost-effective alternative to traditional laboratory techniques.

Furthermore, the COVID-19 pandemic instigated a new focus on rapid molecular diagnostic solutions suitable for point-of-care (POC) use, as the slow turnaround time and centralized processing requirements of current gold-standard options limit their practicality in wide-spread deployment^22,23^. Accordingly, the SmartLid kit was optimized for portability, being easily operable by minimally trained users, stable for room temperature storage, and fully disposable with all required components included and pre-aliquoted.

Here, by using SARS-CoV-2 as a case study, we aim to clinically validate the performance of the SmartLid sample preparation kit for RNA extraction of respiratory viruses, by comparing it with the results obtained by using a gold standard RNA extraction kit (QIAamp Viral RNA Mini Kit, QIAGEN). This innovative approach has the potential to significantly improve access to timely, accurate, and equitable diagnosis of respiratory infections, especially in low-recourse settings and other environments where affordable and portable POC diagnostic capabilities are paramount.

## EXPERIMENTAL SECTION

### SmartLid design and construction

SmartLid(Patent Application Number WO2022180376A1^24^)is a customised magnetic lid that was produced for this study through 3D printing (3D Systems MJP2500Plus) with a UPS Class VI bio-compatible capable material (VisiJet M2S-HT90). Each lid is comprised of two components, a tube closure component (the lid), and a removable magnetic key. The lid contains a recessed bead-collecting surface and incorporates a fluid-wicking member to enhance the removal of liquid to limit carry-over. Furthermore, the lid has two finger tabs for gripping and handling the lid, which not only shield the bead collecting surface from accidental finger or surface contact, but also act as a stand for the lid, allowing the lid to be placed down in between steps (for example, during ethanol evaporation). The magnetic key allows for either collection of the magnetic beads onto the lid (when inserted), or resuspension into each buffer (when removed) without requiring the use of electrical power or special training. SmartLid was designed to work with any single-use Eppendorf tube, to transfer magnetic beads and attached molecules through various preparation steps (lysis-binding, wash, and elution).

Multiple post-processing steps were required to prepare the 3D printed SmartLids for experiments. Initial steps included bulk wax support material removal with steam and fine wax support material removal with heated mineral oil. The SmartLids were then subjected to sonication in 99.95% HPLC grade isopropanol (Sigma Aldrich), followed by sonication in molecular biology grade water (Thermo Scientific), and finally air drying. 5mm by 5mm cylindrical neodymium N42 grade magnets were then glued into the removable key part with cyanoacrylate. Fully assembled SmartLids were subjected to a final UV irradiation step in a PCR cabinet for one hour (30 minutes each side) to limit nucleic acid and nuclease contamination. To reduce waste, magnet keys were reused from batch to batch, until broken, while each closure (lid) part was disposed of after each sample.

### SmartLid sample preparation kit and method

SmartLid can be used with any commercially available magnetic bead-based sample extraction kit by simply prefilling 1.5-2mL flip-cap or screw cap (externally threaded) tubes with prescribed buffers and using SmartLids to transfer the magnetic beads between tubes/steps. However, we designed a single-use POC kit (**Figure 1B**), which includes all required buffers/components, along with a corresponding optimized method, to extract a single sample in less than 5 minutes. Each kit contains: three pre-filled tubes with all necessary buffers in their correct volumes (i.e. tubes 1, 2, and 3, contain magnetic beads with lysis and binding buffer, wash buffer, and elution buffer, respectively), a disposable exact volume pipette, used for transferring the precise amount of inactivated sample (from a swab collection tube for example) into the first tube, the SmartLid, and a plastic tray/package, which doubles as a preparation stand for the process, allowing each reagent tube to stand upright and providing a safe place to rest the SmartLid between steps.

**Figure 1.**
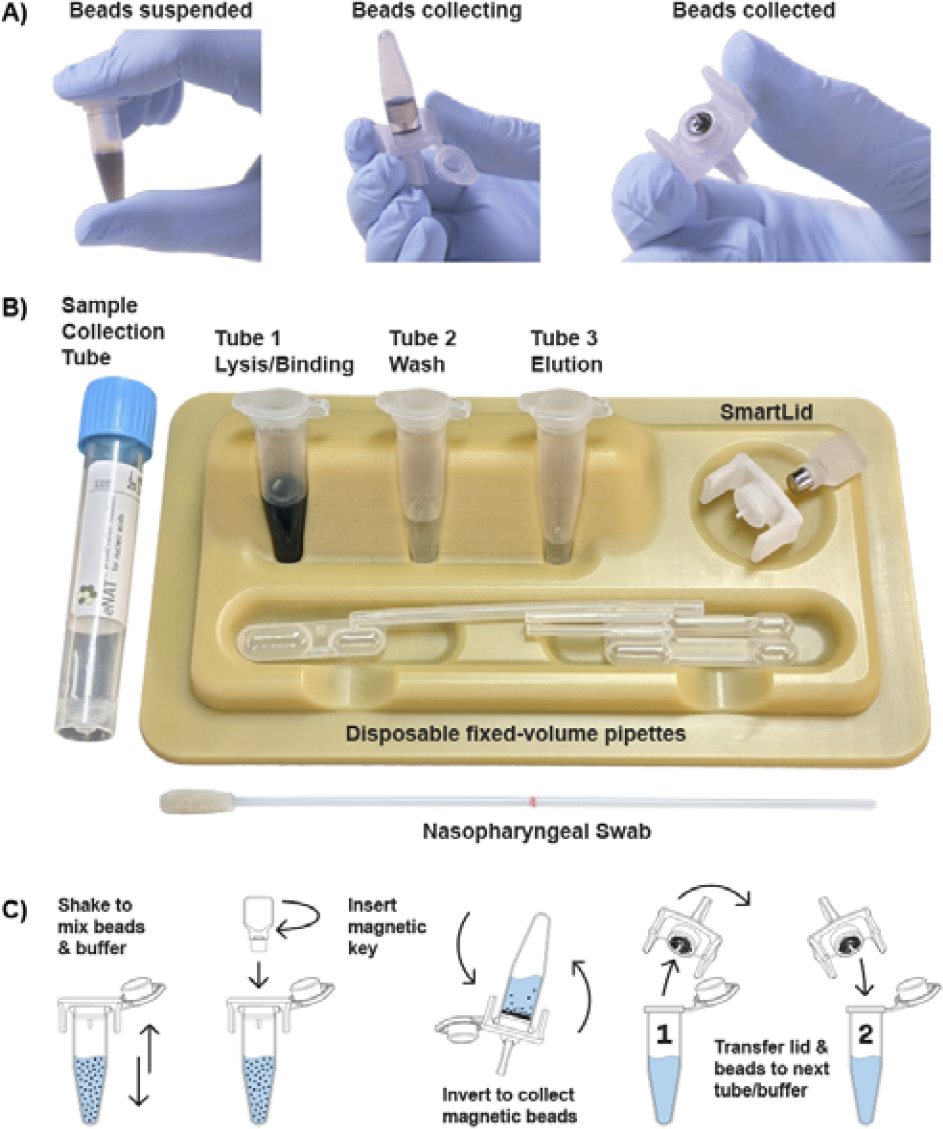
Representative images showing the magnetic bead collection process (A), from full suspension, to partial collection, and full collection. Also shown is the complete single-use SmartLid sample preparation kit (B), and a step-by-step illustrated workflow demonstrating the process to transfer magnetic beads from one tube to another (C).

The detailed method is fully illustrated in supplementary **Figure S1**, ands as follows: A total of 400 μL of sample, stored in an inactivating viral transport medium (Copan neat^®^), is transferred into tube 1 using a fixed-volume pipette. Tube 1 is preloaded with lysis/binding buffer (Turbo Beads LLC) based on guanidinium thiocyanate as the primary component (300μL),99.9% isopropanol (400μL) and silica-based magnetic beads (TurboBeads LLC) suspended in DMSO (20μL). The sample is mixed with the above reagents for 60 seconds through gentle manual shaking or inversion, which binds the present nucleic acids to the surface of the beads. The SmartLid, containing a removable magnetic key, is then used to collect the magnetic beads by inverting the tube (Figure 1A). The SmartLid is then transferred from tube 1 to tube 2, along with the attached magnetic beads and captured nucleic acids. The magnetic key is removed from SmartLid to enable the resuspension of the beads into the wash solution (300μL containing primarily ethanol). The same process of mixing and collecting as above (and illustrated in Figure 1C) is performed, before placing the SmartLid and attached beads down on the provided tray for 30 seconds to allow ethanol evaporation. Finally, the SmartLid and attached nucleic acids are placed into tube 3 (150μLcontaining primarily nuclease-free molecular grade water) for elution of nucleic acids. Once 60 seconds of mixing is complete, the beads are then re-captured onto SmartLid and discarded to leave only eluted nucleic acids and the elution buffer. Purified nucleic acids can then be used for molecular testing or stored at −80 °C until further analysis. The SmartLid sample preparation process can be seen in **Video 1**:https://www.youtube.com/watch?v=KjP3ogoAIXk.

### Respiratory specimens

From October 2020 to January 2021 a total of 406isolates from nasal, nasopharyngeal or buccal swabs were collected at North West London Pathology laboratory (United Kingdom). Clinical samples were obtained previous informed consent from patients and staff undergoing routine SARS-CoV-2 diagnostic testing(including both symptomatic and asymptomatic patients). After initial testing, all samples were stored in 1mL of VTM at −80°C. All residual sample specimens used for this study were inactivated onsite by transferring 500 μL of VTM sample to Copan eNAT at 1:2 ratio prior to shipping the sample to Imperial College London (Hammersmith Campus) for further analysis. Prior to receipt, samples were provided in an anonymised form with no associated data. The study was approved by the Health Research Authority (HRA) and Health and Care Research Wales (HCRW) with NHS Research Ethics Committee (REC) reference 20/HRA/1561.

### RNA extraction with reference method

To characterize the clinical samples, the QIAamp Viral RNA Mini Kit nucleic acid extraction(Catalogue#52904) was used as a reference method for RNA extraction. Sample manipulations (mixing with viral RNA extraction lysis buffer) were performed in a Class II Biosafety Cabinet. Briefly, 140 μL of COPAN eNAT^®^ lysis buffer containing VTM was extracted according to the manufacturer’s instructions^25^. Respiratory swab diluent was added directly to the lysis buffer, vortexed, and incubated for 10 minutes to allow for complete microbial viability inactivation. After a two-step washing process, the RNA was eluted in a total volume of 80 μL. The purified nucleic acids were stored at −80 °C until further analysis.

### RT-qPCR reactions

RNA samples from both the reference and SmartLid methods were tested on a real-time benchtop PCR system (LightCycler 96 Instrument - Roche Life Science) by real-time reverse-transcription PCR (RT-qPCR).For RT-qPCR all experiments, the molecular assay developed by the CDC, defined in “CDC 2019-Novel Coronavirus (2019-nCoV) Real-Time RT-PCR Diagnostic Panel,” was used.26The primers and probes for detecting SARS-CoV-2 were identified from genetic regions belonging to the nucleocapsid (N) gene, encompassing the usage of two primer/probe sets (Supplementary Table S1). An additional primer/probe set was used for amplifying human RNase P gene (RP) in control specimens. The CDC assays for SARS-CoV-2 detection were purchased from Integrated DNA Technologies (Catalogue #10006770).

Briefly, experiments were carried out in duplicates with a final volume of 20 μL per reaction using the assays recommended by the CDC, the N assay (N1 and N2 primer/probe mix) and the RNase P assay (RP primer/probe mix), with the GoTaq Probe 1-Step RT-qPCR (Promega). Each mix contained the following: 10 μL of 2× GoTaq Probe qPCR master mix, 0.4 μL of 50× GoScript RT mix for 1-Step RT-qPCR, 1.5 μL of N1/N2/RNase P assay primer/probe mix, 5 μL of extracted RNA, and enough nuclease-free water to bring the volume to 20 μL. Reactions were performed following the recommendations of the manufacturer: 1 cycle of 15 min at 45°C, 1 cycle of 2 min at 95 °C, and 45 cycles at 95 °C for 3 s and 55°C for 30 s. Reactions were plated in 96-well plates and loaded into a LightCycler 96 real-time PCR system (LC96) (Roche Diagnostics).

A specimen was classified as positive when both the N1 and N2 assays were positive within 40 cycles, which is the lower limit of detection for the CDC RT-qPCR assays. The human RNaseP assay was used as control to reduce false negative rates that can occur due to insufficient number of cells collected and/or poor sample quality. The Ct was calculated by the cycle-threshold method using 0.2 as the fluorescence cutoff value.

### Data analysis and Statistics

The data were analysed using the NumPy and Pandas libraries (Python 3.9), and visualizations were performed with the Matplotlib and seaborn libraries. All the TTP values are reported as mean ± std using built in functions in MATLAB. The analytical performance of the RT-qPCR assays were established in terms of analytical sensitivity, specificity, and accuracy. Sensitivity was calculated as True Positives/(True Positives + False Negatives), Specificity as True Negatives/(True Negatives + False Positives), and Accuracy as (True Positive + True Negative)/total. Correlation was expressed as a linear coefficient of correlation R^2^.

To estimate the number of samples required to be screened, the following formula was used^27^:

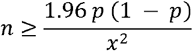

where *p* is the suspected sensitivity, and *x* is the desired margin of error. We define the true-positive rate (sensitivity) as the proportion of SARS-CoV-2 positives which are correctly identified by the SmartLid sample preparation kit compared to the QIAamp Viral RNA Mini Kit. We suspected the sensitivity and specificity of the CDC RT-qPCR assay to be 95% with a desired margin of error of 10%. Under these conditions, the number of required samples is 97 per group. In total, we tested 406 samples (161 positive and 245 negative), exceeding the required sample size per group.

## RESULTS AND DISCUSSION

An ideal POC diagnostic test should fulfil many established characteristics, including affordability, accessibility, being equipment-free and utilizing a user-friendly approach, as established by the World Health Organization REASSURED criteria^28,29,18,30^. Based on these guidelines, current gold standard laboratory-based sample preparation methods have limited use in the future of viral diagnostics at the POC. They are not only space-demanding and rely on complicated and sensitive procedures, but also require bulky benchtop equipment and extensive training to complete. In contrast, our solution has a simple and user-friendly workflow, not requiring any specialized scientific training, while also remaining electricity and micro-pipette free. Furthermore, we estimate the cost of RNA isolation using the SmartLid sample prep kit to be significantly lower than competing gold standard methods, especially when considering the lack of infrastructure and hardware required.

Protocol time in particular is one of the main functional requirements for the development of innovative diagnostic tools, with more rapid and efficient results able to yield more timely and appropriate treatment decisions. Extraction and purification of nucleic acids is one of, if not the most time-consuming steps in the diagnostic process. In fact, reference methods for SARS-CoV-2 RNA extraction often take at least 25 minutes, but frequently anywhere from 45-60 minutes, to complete^31^. In contrast, as shown in **Video 1**, total RNA extraction utilizing SmartLid takes less than 5 minutes and can be performed at the POC, reducing additional delays in the diagnostic process by avoiding the need for transporting samples to a centralized laboratory.

Contributing to SmartLid’s POC suitability, the kit packaging was designed to act as a sample processing stand, eliminating the need for any externally sourced racks and minimizing the possibility of contamination, with convenient locations for all components during the procedure. In fact, there is no requirement even for a bench or table, as the process can be performed on any surface, including the user’s lap while sitting. Furthermore, all buffers and kit components were chosen for their room temperature stability, removing the requirement for cold chain storage and shipment. In terms of size, a complete disposable single-use kit, including all essential buffers, plastics, packaging, and SmartLid can easily fit in the palm of a hand.

### Validation and clinical performance of the SmartLid sample preparation kit

To ensure that the SmartLid purification method maintained high sensitivity and specificity, we validated the methodology by comparing its performance against the gold standard for viral RNA extraction, the Viral RNA Mini Kit (QIAGEN).RT-qPCR using primers recommended by the CDC (**Supplementary Table S1**) was performed with residual clinical samples from patients and personnel who were subjected to standard SARS-CoV-2 diagnostic testing. The nucleocapsid viral proteins N1 and N2 were amplified as viral targets, and RNase P was also amplified as a human gene control. We analysed 406 clinical isolates: 161 were determined as SARS-CoV-2 positive and 245 as SARS-CoV-2 negative according to RT-qPCR using RNA extracted with the reference method. Estimated RNA concentration for positive specimens ranged from 10^9^ to 10^2^ copies per mL of storage buffer, covering all clinically relevant ranges of viral loads, as shown in **Supplementary Figure S2**. RT-qPCR results obtained with the N1 assay were used to classify the samples into 5 categories according to RNA concentration (high, upper-medium, lower-medium, low, and negative).Specific concentration ranges for these categories are shown in **Supplementary Table S2**.

A total of153 samples were detected by N1 and N2, and 406 samples by RNase P according to RT-qPCR results with RNA extracted using the SmartLid method. Eight false negatives and one false positive were obtained when compared to the QIAGEN method, which translates to an overall sensitivity of 95.03% (95% CI: 90.44%–97.83%) and a specificity of 99.59% (95% CI: 97.76%–99.99%), with a positive agreement of 97.79% (95% CI: 95.84%–98.98%). Determination of the sensitivity, specificity and accuracy across the 5 sample categories is shown in **Table 1**.

**Table 1.** SmartLid method performance based on sample category.

| Categories<br>(range Ct)<sup>1</sup> | True<br>Positives | True<br>Negatives | False<br>Positives | False<br>Negatives | Sensitivity<br>(%) | Specificity<br>(%) | Accuracy<br>(%) |
| --- | --- | --- | --- | --- | --- | --- | --- |
| High (14-20.9) | 48 | - | - | 0 | 100 | - | 99.6 |
| Upper-medium (21-24.9) | 40 | - | - | 0 | 100 | - | 99.6 |
| Lower-medium (25-30.9) | 38 | - | - | 0 | 100 | - | 99.6 |
| Low (31-40) | 26 | - | - | 8* | 76.5 | - | 96.8 |
| Negative (not detected) | - | 244 | 1 | - | - | 99.6 | - |
| Total | 153 | 244 | 1 | 8 | 95.0 | 99.6 | 97.8 |

<sup>1</sup>Categories are defined based on the QIAGEN method and CDC RT-qPCR N1 assay values. \*Note: all false-negative results were samples with an average N1 Ct value of 34.38, which corresponds to a copy number lower than $1.4 \times 10^3$ copies/ml of lysis buffer

Although SmartLid achieved high sensitivity and specificity when compared with the gold standard, eight positive samples with low viral loads were not detected. All had a Ct value greater than 31.7 in N1 and 35.5 in N2, with an average Ct value of 34.38 and 37.70 respectively. Furthermore, only three out of the eight false negative results showed a false negative result for both the N1 and N2 genes. Finally, the single false positive result was also positive in N1 for the reference method, potentially indicating a correct classification of a sample with extremely low viral load by SmartLid, which was missed by the reference method.

### Analytical sensitivity of the SmartLid sample preparation kit

The analytical sensitivity for the SmartLid method was addressed by comparing the Ct values obtained for the N1, N2 and RNase P targets with the reference method (**Figure 2**).The mean Ct values for N1, N2 and RNaseP with the QIAGEN RNA extraction method were 25.66 ± 6.54, 26.96 ±6.91 and 29.65 ± 2.90, respectively. For the SmartLid Kit, mean Ct values were 28.11 ±5.39, 29.40 ± 6.15, 30.43 ± 3.25, respectively (full data set is shown in **Supplementary Information_2 file**). Overall, we found that Ct values for N1 and N2 were statistically significantly higher than those obtained with the QIAGEN kit (*p* = 3.778 × 10^−5^). No significant differences were observed for RNaseP (*p* = 2.364 × 10^−2^).As mentioned above, however, it is likely that the larger elution volume used for the SmartLid method played a role, accounting for an estimated 1 Ct difference alone due to the dilution factor.

**Figure 2.**
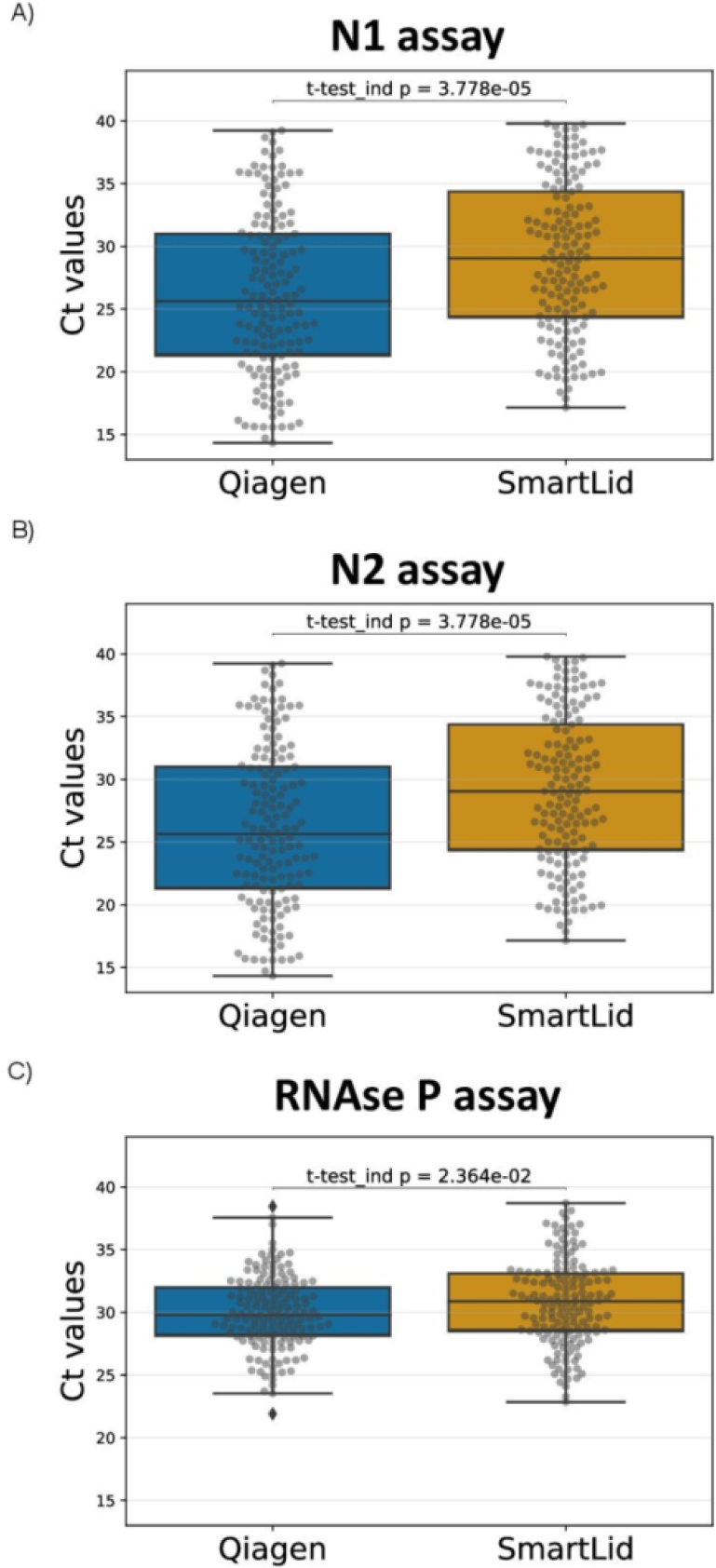
Comparison of Ct value distribution between QIAGEN and SmartLid methods for CDC qRT-PCR N1 (A), N2 (B) and RNase P (C) genes. Data expressed as box and whisker plots, showing median (horizontal line), boxes epresenting the 25th to 75th percentiles, whiskers representing minimum and maximum values.

Technical differences in the extraction and purification process could explain these results, such as eluting in a volume roughly 2 times larger than the reference method (diluting our end product accordingly), fewer total steps, and a significantly shorter protocol time. For example, the reference method took more almost ten times longer to complete than the SmartLid method for a single sample, and more than three times longer even when processing multiple samples (12-24) simultaneously. Nonetheless, one should consider that samples amplifying at a cycle over 30 (Ct 30-40) are typically associated with non-infectious patients32,33, and that the sensitivity of our method increased to 100% with samples in the lower-medium to high viral load categories (Ct <31) (**Table 1)**.

To further evaluate the performance of the SmartLid method, the correlation between Ct values obtained by both methods was computed by applying linear regression (**Figure 3**). The SmartLid and QIAGEN methods showed a correlation for the RT-qPCR N1, N2 and RNaseP assays of R^2^=0.88, R^2^=0.90 and R^2^=0.71, respectively.

**Figure 3.**
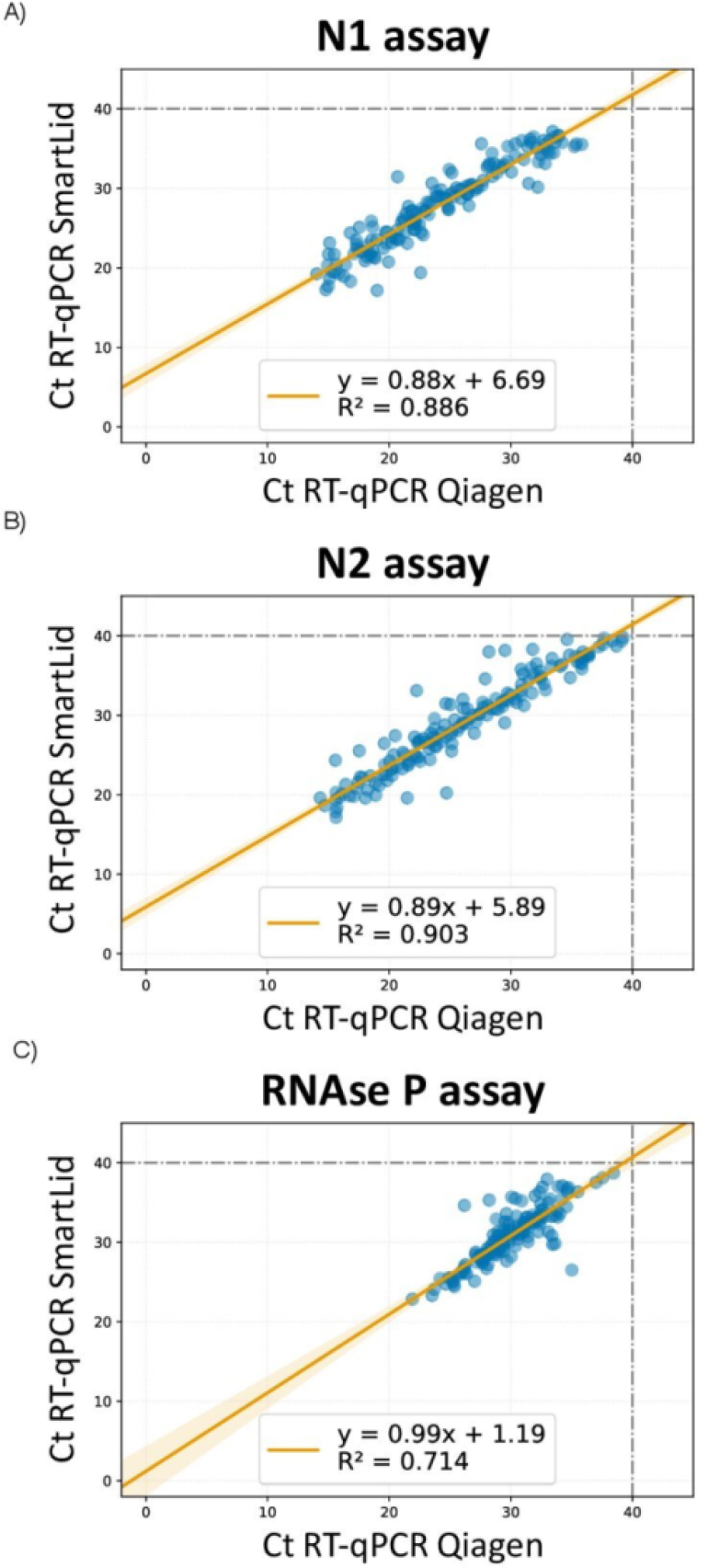
Correlation between cycle threshold (Ct) values of CDC RT-qPCR *N1* (A), *N2* (B) and *RNase P* (C) genes using RNA purified with SmartLid and QIAGEN methods.

Sample extraction is a critical step in diagnostic testing procedures, and the demand for efficient and reliable methods, particularly within POC settings in the absence of laboratory infrastructure, has only been heightened as a result of the SARS-CoV-2 pandemic^34^. Various abbreviated sample extraction methods have been developed to address this need, including at one extreme direct nucleic acid amplification in unprocessed and unpurified samples, which offers speed and scalability advantages^35–37^. However, these methods often exhibit lower sensitivity and accuracy compared to processes with more emphasis on thorough sample extraction prior to amplification. Sample extraction and purification is important because it removes unwanted materials and contaminants from the sample, such as proteins and lipids, leading to a cleaner and more concentrated sample for downstream analysis with less inhibition.

Our approach offers a unique balanced solution at the intersection of speed and performance. Despite the fact that mean Ct values for the SmartLid extracted samples were higher than the reference values for the N1 and N2 target genes, the SmartLid method’s overall performance combined with its simple and quick procedure implies significant benefits over gold standard sample preparation methods.

## CONCLUSIONS

In this study, we described a unique, electricity-free, portable sample extraction method, based on a custom magnetic lid (SmartLid). We clinically evaluated our approach using 406 residual samples from patients tested for SARS-CoV-2 and demonstrated its efficacy compared to the QIAamp Viral RNA reference method from QIAGEN. Results revealed comparable performance between our sample extraction method and the QIAGEN procedure while taking less than 5 minutes from start to finish and requiring no benchtop equipment and no specialize training. This is in stark contrast to QIAGEN’s method which takes 45-60 minutes to complete, requires expensive and bulky benchtop centrifuges, vortex mixers, and micropipettes, as well as necessitates personnel with advanced training. Such properties make SmartLid an exciting option for accelerating the advancement of molecular diagnostic capabilities in areas where funding and access to laboratory infrastructure is limited, but accurate testing is still of upmost importance. Therefore, SmartLid represents a step-change in POC sample extraction, enabling rapid and cost-effective diagnosis of COVID-19 as well as other infectious diseases.

Since efficient and reproducible sample extraction is a prerequisite for a robust and sensitive quantitative molecular workflow, the SmartLid technology and method is currently undergoing a number of improvements. For example, we are adapting the protocol and buffers for the extraction of not only viruses, but also bacteria, and ensuring that SmartLid performs optimally with a range of sample types, including nasal, nasopharyngeal, and buccal swabs, saliva, non-inactivating/lysis transport media, blood, serum, plasma, stool, waste-water, and more. Furthermore, since the kit is intended to be used in POC settings, we are aiming to reduce the significant quantity of plastic waste that may be generated with each component being disposable. To offset this, we are exploring iterations of the packaging/preparation stand made with 100% recycled cardboard, leaving only the SmartLid, buffer tubes, and disposable exact volume pipette as plastic waste, as well as developing a bulk-packed version. Additionally, the magnetic key can easily be reused from sample to sample, as it poses minimal risk of cross-contamination due to its limited exposure to the sample. Other improvements are being considered with respect to long term storage and shipping. We have observed that the ethanol in tube 2 can experience significant evaporation (20-30% of the volume lost after 3-4 months) when stored at room temperature. Moreover, ethanol (in concentrations above 10%) is considered a flammability risk according to most international shipping guidelines. To mitigate both of these concerns, we are investigating the use of different types of tubes, both those with more secure flip-top closures (Safe-Lock from Eppendorf^®^ for example) and gasketed screw-top closures (CRYOVIAL^®^ from Simport Scientific), some of which have shown a reduction in evaporation loss and would decrease the chance of leakage during shipping. However, all of the above improvements and further investigations are out of the scope of this manuscript.

Finally, with the goal of developing a truly accessible sample-to-result molecular diagnostic test, we envision the coupling of a simple and frugal detection method with SmartLid, which will usher in the next generation of truly POC molecular diagnostic solutions for resource-limited settings.

## Data Availability

All data produced in the present study are available upon reasonable request to the authors

## ASSOCIATED CONTENT

### Supporting Information

The Supporting Information is available free of charge on the ACS Publications website.

**Supplementary_Information_1 file**: CDC RT-qPCR panel: primers and probes (**Table S1**),clinical sample characterization by CDC RT-qPCR, based on N1 CDC assay (**Table S2**), Schematic representation of sample workflow from swabbed sample to eluted product using SmartLid technology (**Figure S1**), Estimated SARS-CoV-2 RNA concentration (copies/500μL lysis buffer) across positive samples (**Figure S2**)(PDF). Complete residual clinical sample testing data (EXCEL).

**Supplementary_Information_2 file:** Results of the clinical test data.

## AUTHOR INFORMATION

### Author Contributions

Study concept and design: IP,MLC, JRM. Acquisition, analysis, or interpretation of data: All authors. Drafting the manuscript:IP, MLC, LM, JRM. Critical revision of the manuscript: All authors. The manuscript was written through the contributions of all authors. All authors have given approval to the final version of the manuscript.

### Declaration of competing interest

IP, MLC, KMC, KTM, NM, PG and JRM have financial interest on ProtonDx Ltd, which currently has an exclusive license to the SmartLid intellectual property and trademark. All authors declare that they have no other conflict of interest related to this work.All the authors declare that they do not have any other known competing financial interests or personal relationships that could have appeared to influence the work reported in this paper.

### Funding

This work was supported by the Department of Health and Social Care-funded Centre for Antimicrobial Optimisation (CAMO) at Imperial College London; The Imperial College 3M/Global Giving Foundation COVID-19 Research Award; and the Imperial COVID-19 Research Fund. IP acknowledges the NIH Research award (WMNP_P61895) at Imperial College London. AH, PG and JRM are affiliated with the NIHR Health Protection Research Unit (HPRU) in Healthcare Associated Infections and Antimicrobial Resistance at Imperial College London in partnership with the UK Health Security Agency, in collaboration with, Imperial Healthcare Partners, the University of Cambridge and the University of Warwick. The views expressed in this publication are those of the authors and not necessarily those of the NHS, the National Institute for Health Research, the Department of Health and Social Care, or the UK Health Security Agency. AH is a National Institute for Health Research (NIHR) Senior Investigator.

## Notes

### Author Declarations

The study was approved by the Health Research Authority (HRA) and Health and Care Research Wales (HCRW) with NHS Research Ethics Committee (REC) reference 20/HRA/1561.

