## Supplementary Figures and Tables for "A Rapid, Portable, and Electricity-Free Sample Extraction Method for Enhanced Molecular Diagnostics in Resource-Limited Settings"

**Supplementary materials**

**Supplementary tables**

**Table S1.** CDC RT-qPCR panel: primers and probes

| **Name** | **Description** | **Oligonucleotide sequence (5'→3')** | **Label** | **Final conc.** |
| --- | --- | --- | --- | --- |
| 2019-nCoV_N1-F | 2019-nCoV_N1 Forward Primer | 5’-GAC CCC AAA ATC AGC GAA AT-3’ | None | 500nM |
| 2019-nCoV_N1-R | 2019-nCoV_N1 Reverse Primer | 5’-TCT GGT TAC TGC CAG TTG AAT CTG-3’ | None | 500nM |
| 2019-nCoV_N1-P | 2019-nCoV_N1 Probe | 5’-FAM-ACC CCG CAT TAC GTT TGG TGG ACC-BHQ1-3’ | FAM, BHQ-1 | 125nM |
| 2019-nCoV_N2-F | 2019-nCoV_N2 Forward Primer | 5’-TTA CAA ACA TTG GCC GCA AA-3’ | None | 500nM |
| 2019-nCoV_N2-R | 2019-nCoV_N2 Reverse Primer | 5’-GCG CGA CAT TCC GAA GAA-3’ | None | 500nM |
| 2019-nCoV_N2-P | 2019-nCoV_N2 Probe | 5’-FAM-ACA ATT TGC CCC CAG CGC TTC AG-BHQ1-3’ | FAM, BHQ-1 | 125nM |
| RP-F | RNAse P Forward Primer | AGA TTT GGA CCT GCG AGC G | None | 500nM |
| RP-R | RNAse P Reverse Primer | GAG CGG CTG TCT CCA CAA GT | None | 500nM |
| RP-P | RNAse P probe | FAM – TTC TGA CCT GAA GGC TCT GCG CG – BHQ-1 | FAM, BHQ-1 | 125nM |

| **Categories^a^** | **RT-qPCR *C_t_* range**  **N1 assay (cycles)** | **RNA concentration**  **(copies/reaction)^b^** | **RNA concentration**  **(copies/mL of Lysis Buffer)^c^** | **Samples per category**  **(*n*)** |
| --- | --- | --- | --- | --- |
| High | 14 – 20.9 | 1 x 10^7^ – 1 x 10^5^ | 2 x 10^9^ – 1 x 10^7^ | 48 |
| Upper-medium | 21 – 24.9 | 1 x 10^5^ – 8 x 10^3^ | 1 x 10^7^ – 1 x 10^6^ | 40 |
| Lower-medium | 25 – 30.9 | 8 x 10^3^ – 1 x 10^2^ | 1 x 10^6^ – 1 x 10^4^ | 38 |
| Low | 31 – 37.6 | 1 x 10^2^ – 1 x 10^0^ | 1 x 10^4^ – 1 x 10^2^ | 35 |
| Negative | Not detected | – | – | 245 |
| Total | - |  |  | 406 |

**Table S2.** Clinical sample characterization by CDC RT-qPCR, based on N1 CDC assay

^a^All samples tested positive by the RNAse P assay. ^b^Estimated concentration (copies/reaction) based on RT-qPCR standard curve y = −3.30x + 37.98. ^c^Estimated concentration (copies/mL of Lysis Buffer) considering 140μL of sample input and 80μL eluted volume.

**Supplementary figures**

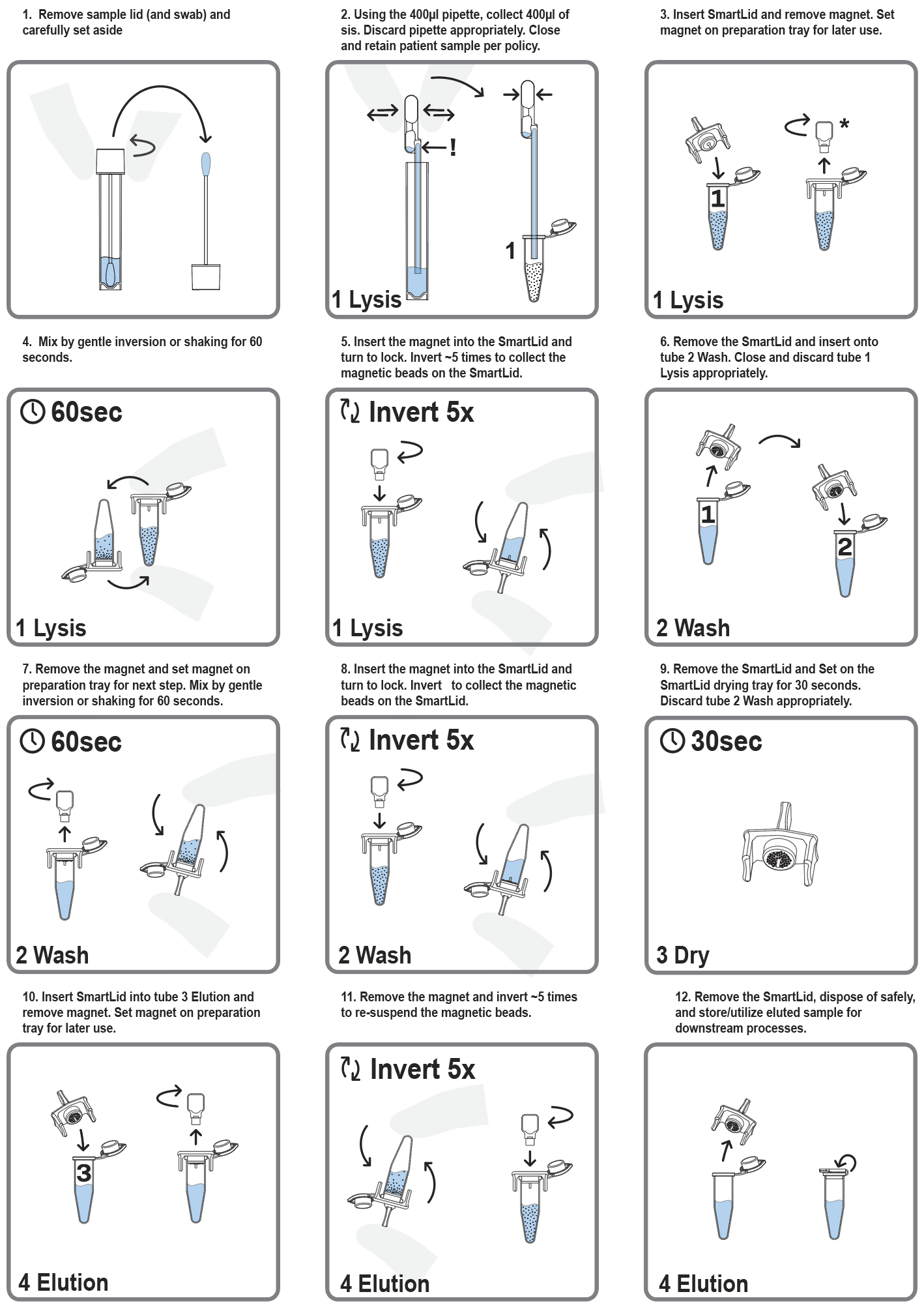

**Figure S1.** Schematic representation of sample workflow from swabbed sample to eluted product using SmartLid technology.

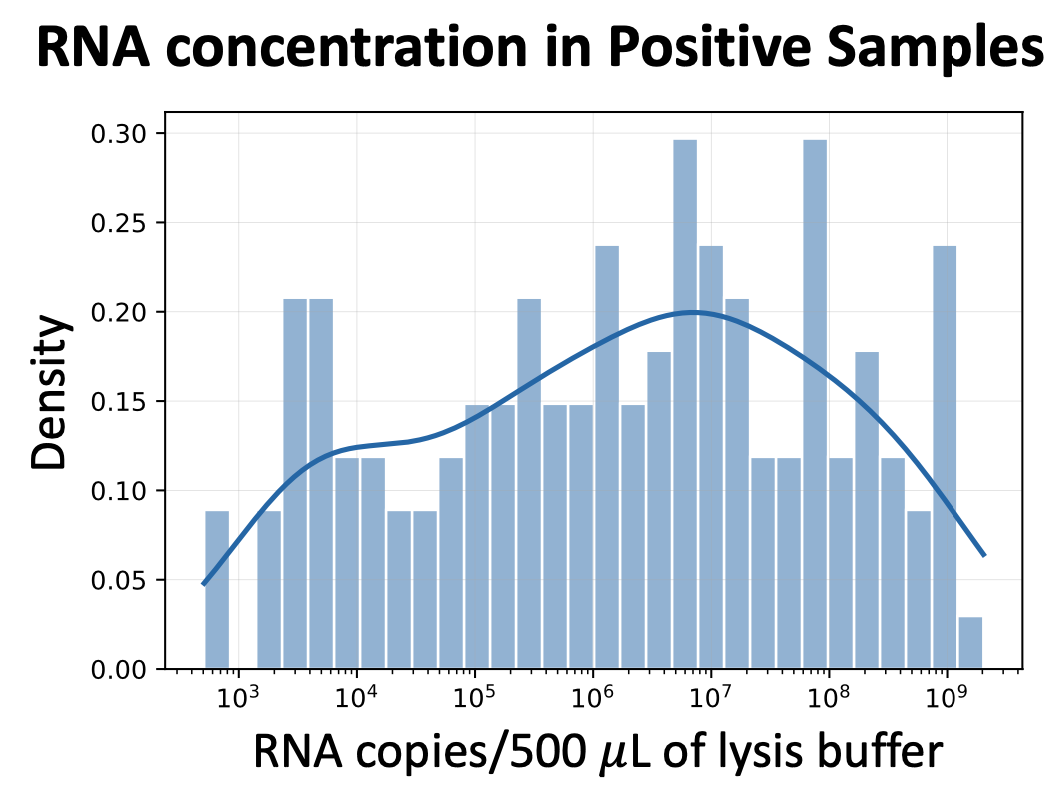

**Figure S2.** Estimated SARS-CoV-2 RNA concentration (copies/500 µL lysis buffer) across positive samples.
